## Supplementary files for "Effectiveness of early versus delayed rehabilitation following rotator cuff repair: systematic review and meta-analyses"

**Supplementary file 1.** Search strategy

1. Shoulder/ or Shoulder Injuries/ or Shoulder Joint/ or Shoulder Pain/

2. Rotator Cuff/

3. "Physical and Rehabilitation Medicine"/ or Rehabilitation/

4. Physical Therapy Modalities/ or Exercise Therapy/

5. Physiotherapy.mp. [mp=title, abstract, original title, name of substance word, subject heading word, floating sub-heading word, keyword heading word, organism supplementary concept word, protocol supplementary concept word, rare disease supplementary concept word, unique identifier, synonyms]

6. 1 or 2

7. 3 or 4 or 5

8. 6 and 7

9. randomi*ed controlled trial.mp. [mp=title, abstract, original title, name of substance word, subject heading word, floating sub-heading word, keyword heading word, organism supplementary concept word, protocol supplementary concept word, rare disease supplementary concept word, unique identifier, synonyms]

10. Clinical Trial/ or Controlled Clinical Trial/ or Pragmatic Clinical Trial/

11. 9 or 10

12. 8 and 11

**Supplementary file 2.** Characteristics of the included RCTs.

| **Author (year)** | **Country** | **No. of patients randomised**  **E/D – M/F** | **Age**  **(years) E/D** | **Tear characteristics** | **Surgery characteristics** | **Outcomes** |
| --- | --- | --- | --- | --- | --- | --- |
| Arndt, Clavert (1) | France | 49/43 – 58/34 | 55.3 | Non-retracted isolated tears of supraspinatus; partial-thickness: 24%, full-thickness: 76% | 5 surgeons;  59% single row, 41% double row; LHB tenotomy: 65%, LHB tenodesys:11%; acromioplasty: 91% | CM, cuff integrity (arthrogram, CT or arthro-MRI), ROM |
| Cuff and Pupello (2) | USA | 33/35 – 38/30 | 63/63.5 | Supraspinatus; full-thickness; crescent-shaped | Transosseous suture bridge | ASES, cuff integrity (US), ROM, SST |
| De Roo, Muermans (3) | Belgium | 79/51 – 89/41 | 65.1/64.6 | Small to large; full-thickness | Single or double row; acromioplasty | CM, cuff integrity (US) ROM, SPADI, SST, strength (dynamometer), UCLA |
| Duzgun, Baltaci (4) | Turkey | 13/16 – 3/26 | 55.8/56.6 | Medium and large | NA | DASH, pain (VAS), ROM |

***Continue***

**Supplementary file 2 (continue).** Characteristics of the included RCTs.

| **Author (year)** | **Country** | **No. of patients randomised**  **E/D – M/F** | **Age**  **(years) E/D** | **Tear characteristics** | **Surgery characteristics** | **Outcomes** |
| --- | --- | --- | --- | --- | --- | --- |
| Duzgun, Baltaci (5) | Turkey | 20/22 – 6/34 | 57.6/57.2 | Medium and large | NA | ROM |
| Fawzy, Rizk Mohamed (6) | Egypt | 86/86 – 90/74 | 57.8/57 | Small to Medium sized; full-thickness | Single-row and subacromial decompression; no biceps procedures | ASES, CM, cuff integrity (MRI), pain (VAS) ROM |
| Jenssen, Lundgreen (7) | Norway | 60/60 – 69/49 | 56/55 | Supraspinatus or upper infraspinatus; small to medium sized; full-thickness | Different surgeons; single-row using 1 or 2 triple-loaded suture anchors; subacromial decompression; LHB tenotomy/tenodesis (no difference between groups ); ACJ resections (E: 8% vs. D: 23%) | CM, cuff integrity (MRI), ROM, WORC |
| Keener, Galatz (8) | USA | 67/62 – 73/51 | 55.8/54.8 | Supraspinatus and/or infraspinatus; Small and medium; full-thickness | 3 surgeons; double row transosseous; acromioplasty; LHB tenodesis or tenotomy | ASES, CM, cuff integrity (US), pain (VAS) ROM, SST, strength |

**Supplementary file 2.** Characteristics of the included RCTs.

| **Author (year)** | **Country** | **No. of patients randomised**  **E/D – M/F** | **Age**  **(years) E/D** | **Tear characteristics** | **Surgery characteristics** | **Outcomes** |
| --- | --- | --- | --- | --- | --- | --- |
| Kim, Chung (9) | Korea | 60/57 - 44/61 | 60/60 | Small and medium; full-thickness | Different surgeons; single row: 17, double row: 2, suture bridge: 86; acromioplasty | ASES, CM, cuff integrity (US,MRI or CT), pain (VAS) ROM, SST |
| Kjær (10) | Denmark | 41/41 – 64/28 | 59/61 | Supraspinatus involved: E: 100% vs D:100%  Infraspinatus involved: E:26% vs D:36%  Subscapularis involved: E:7.3% vs D:17.1% | NA | Cuff integrity (US), DASH, pain (NRS), ROM, strength (dynamometer), WORC |
| Klintberg, Gunnarsson (11) | Sweden | 9/9 – 9/5 | 55 | Full-thickness | NA | CM, FIS, pain (VAS) ROM, strength (isokinetics) |
| ***Continue*** |  |  |  |  |  |  |

**Supplementary file 2.** Characteristics of the included RCTs.

| **Author (year)** | **Country** | **No. of patients randomised**  **E/D – M/F** | **Age**  **(years) E/D** | **Tear characteristics** | **Surgery characteristics** | **Outcomes** |
| --- | --- | --- | --- | --- | --- | --- |
| Koh, Lim (12) | Korea | 47/53 – 44/44 | 60.1/59.5 | Postero-superior; medium; full-thickness;  2-4 cm | Single row, acromioplasty, capsular release | ASES, CM, cuff integrity (MRI), VAS |
| Lee, Cho (13) | Korea | 43/42 – 41/23 | 54.5/55.2 | Medium: 41, large: 45; full-thickness | One surgeon; single row; patients who need LHB, acromion and/or clavicle procedures were excluded | Cuff integrity (MRI), ROM, strength (dynamometer), UCLA, VAS |
| Littlewood, Bateman (14) | UK | 37/36 – 42/31 | 60.6/65.4 | All sizes included: E:2.96 cm vs D:2.5 cm  Supraspinatus involved: E: 28 vs D:30  Infraspinatus involved: E:7 vs D:6  Subscapularis involved: E:1% vs D:6% | Eight surgeons;  complete repairs: E: 17 vs D:30  partial repairs:  E: 6 vs D:0 | Cuff integrity (US), EQ-5D-5L, OSS, SPADI |
| ***Continue*** |  |  |  |  |  |  |

**Supplementary file 2.** Characteristics of the included RCTs.

| **Author (year)** | **Country** | **No. of patients randomised**  **E/D – M/F** | **Age**  **(years) E/D** | **Tear characteristics** | **Surgery characteristics** | **Outcomes** |
| --- | --- | --- | --- | --- | --- | --- |
| Mazzocca, Arciero (15) | USA | 36/37 – 40/18 | 54/55 | Supraspinatus; full-thickness | Single surgeon; transosseous equivalent; three to four anchors; subacromial decompression; LHB tenodesis | ASES, CM, cuff integrity (MRI), ROM, SANE, SST, WORC |
| Oyarzún, Poblete (16) | Chile | 15/15 – 22/8 | NA | Supraspinatus | NA | Pain (VAS), ROM |
| Raschhofer, Poulios (17) | Austria | 14/16 – 19/10 | 56.3/59.5 | Medium sized; full-thickness | Single-row; subacromial decompression; biceps tenotomy and ACJ resection | CM, DASH, pain (VAS), ROM, strength (dynamometer) |
| ***Continue*** |  |  |  |  |  |  |

**Supplementary file 2.** Characteristics of the included RCTs.

| **Author (year)** | **Country** | **No. of patients randomised**  **E/D – M/F** | **Age**  **(years) E/D** | **Tear characteristics** | **Surgery characteristics** | **Outcomes** |
| --- | --- | --- | --- | --- | --- | --- |
| Sheps, Bouliane (18) | Canada | 97/92 – 115/74 | 55.4/54.9 | All tear sizes included.  Single tendon involvement: E:79% vs D: 84%; tear sizes: Small: E:28% vs D: 26%  Medium: E: 51% vs D: 56%  Large: E: 21% vs D:18%;  full-thickness | Multiple surgeons; mini-open method | Cuff integrity (NA), pain (VAS), ROM, strength (tensiometer), WORC |
| ***Continue*** |  |  |  |  |  |  |

**Supplementary file 2.** Characteristics of the included RCTs.

| **Author (year)** | **Country** | **No. of patients randomised**  **E/D – M/F** | **Age**  **(years) E/D** | **Tear characteristics** | **Surgery characteristics** | **Outcomes** |
| --- | --- | --- | --- | --- | --- | --- |
| Sheps, Silveira (19) | Canada | 103/103 – 131/75 | 55.5/56.2 | All tear sizes included.  Mean length of tear AP: E: 2.1 cm vs D: 2.1 cm  Mean length of tear ML: E: 1.9 cm vs D: 1.9 cm | Multiple surgeons; arthroscopic method. Single row: E: 10.7% vs D:10.7%  Double row/transosseous: E: 89.3% vs D: 89.3%  Tenodesis: E: 45% vs D: 44.1  Acromioplasty: E: 75.7% vs D:77.5%  ACJ excision: E:15.5% vs D:9.7% | Cuff integrity (US), pain (VAS), ROM, SF-36, strength, (dynamometer), WORC |
| Tirefort, Schwitzguebel (20) | Switzerland | 40/40 – 37/43 | 54.7/53.5 | Isolated superior full thickness tear; small to medium sized | Double row suture anchors.  Tenodesis: E:53% vs D: 65%  Tenotomy: E: 38% vs D: 33%  Acromioplasty: E:98% vs D: 88% ACJ resection: E:35% vs D: 33% | ASES, cuff integrity (US), pain (VAS), ROM, SANE |

ACJ: acromioclavicular joint, AP: anteroposterior, ASES: American Shoulder and Elbow Surgeons, CT: Computed Tomography, CM: Constant-Murley Score, E/D: Early/Delayed, FIS: Functional Index of the Shoulder, LHB: Long Head of Biceps, MRI: Magnetic Resonance Imaging, M/F: Male/Female, ML: mediolateral, NA: Not Available, OSS: Oxford Shoulder Score, ROM: Range Of Motion, RCT: Rotator Cuff Tear, SANE: Single Assessment Numeric Evaluation score, SPADI: Shoulder Pain and Disability Index, SST: Simple Shoulder Test Score, US: Ultrasound, UCLA: University of California Los Angeles, VAS: Visual Analogue Scale, WORC: Western Ontario Rotator Cuff index.

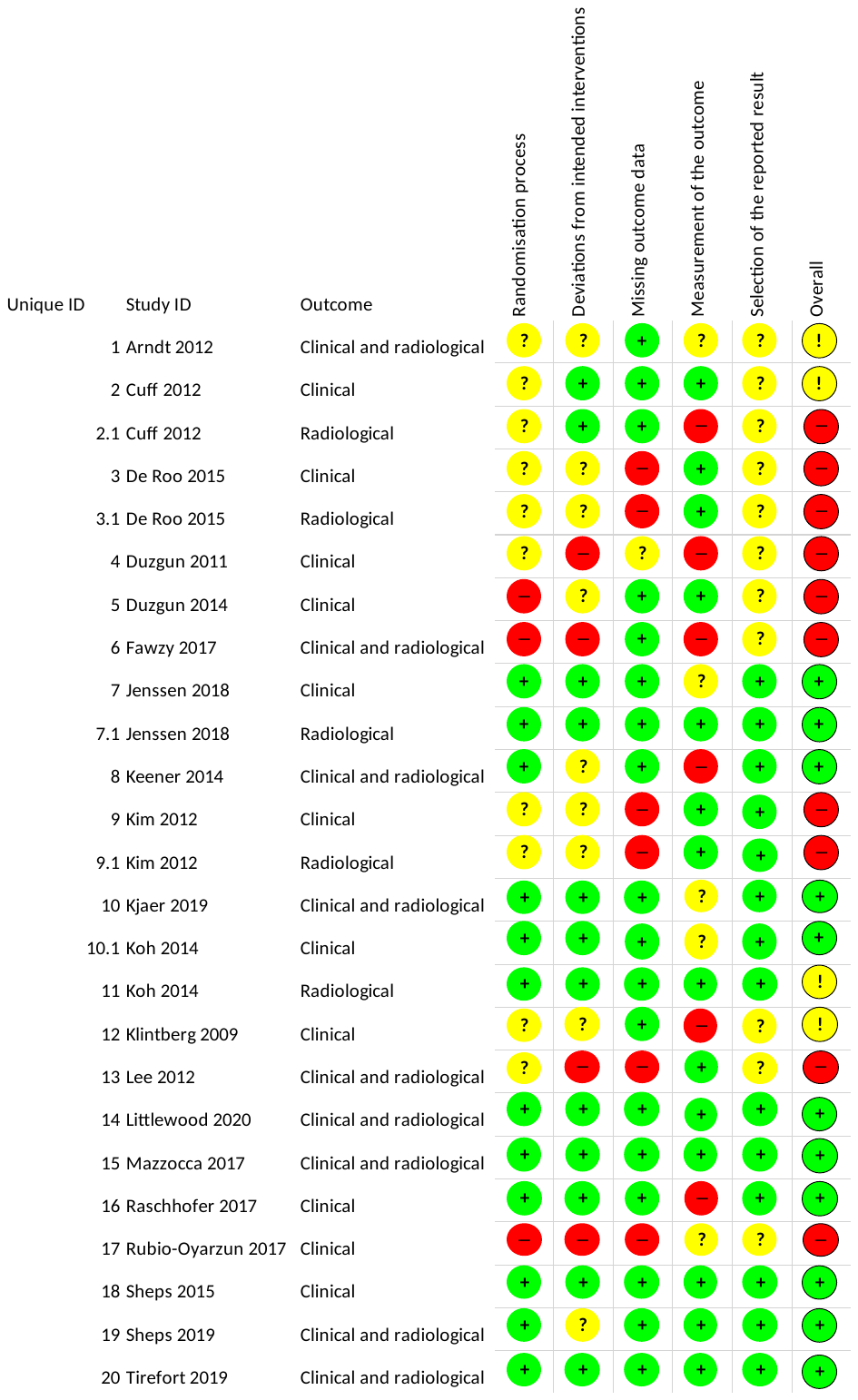
**Supplementary file 3.** Risk of bias by RCT.

**Supplementary file 4.** Characteristics of the rehabilitation programmes.

| **Author (year)** | **Early Rehabilitation** | **Delayed Rehabilitation** |
| --- | --- | --- |
| Arndt, Clavert (1) | **IP:** Sling for 6 weeks  **First day postoperative-week 6:** Pendulum exercise **+** manual shoulder PROM exercises **+** CPM (3-5x per week)  **Week 6-4 Months:** Shoulder AROM exercises  **From 4 months:** Strengthening exercises | **IP:** Sling for 6 weeks  **Week 0-6:** Immobilisation **+** Pendulum exercise  **Week 6-4 Months:** Shoulder AROM exercises  **From 4 months:** Strengthening exercises |
| Cuff and Pupello (2) | **IP:** Shoulder immobiliser for 6 weeks  **Weeks 0-3:** Started in the second day post-surgery. Pendulum exercise **+** shoulder PROM exercises for flexion and external rotation **+** elbow, wrist and hand AROM exercises  **Weeks 4-6:** Similar to week 0-3 **+** progressing ROM **+** AROM elbow, wrist and hand  **Weeks 6-10:** Shoulder AAROM exercises  **Weeks 10-12:** Shoulder AAROM **+** AROM exercises  **From week 12:** Strengthening  Face-to-face sessions 3x per week | **IP:** Shoulder immobiliser for 6 weeks  **Weeks 0-3:** Pendulum exercise 3x daily for 5 minutes **+** elbow, wrist and hand AROM exercises  **Weeks 4-6:** Pendulum exercise 3x daily for 5 minutes **+** elbow, wrist and hand AROM exercises  **Weeks 6-10:** Shoulder PROM exercises **+ week 7** Shoulder AROM exercises 1x per week  **Weeks 10-12:** Shoulder AAROM **+** AROM exercises  **From week 12:** Strengthening |
| ***Continue*** |  |  |

**Supplementary file 4.** Characteristics of the rehabilitation programmes.

| **Author (year)** | **Early Rehabilitation** | **Delayed Rehabilitation** |
| --- | --- | --- |
| De Roo, Muermans (3) | **IP:** Brace with abduction pillow (30°) for 4 weeks during day and night **+** 2 more weeks only at night  **First day postoperative – week 5:** Pendulum exercise (3x per day, max 10 minutes, 20 cm diameter) **+** shoulder PROM for flexion, abduction, internal and external rotation **+** scapular mobilization (5 days pw)  **Weeks 5-8:** Specific capsular glenohumeral exercises **+** shoulder AAROM exercises  **From week 8:** Strengthening | **IP:** Brace with abduction pillow (30°) for 4 weeks during day and night **+** 2 more weeks only at night  **Weeks 1-4:** Pendulum exercise (3x per day, max 10 minutes, 20 cm diameter)  **Weeks 5:** Gradual shoulder PROM mobilization  **From week 6:** Similar to early mobilisation group; no further details available |
| ***Continue*** |  |  |

**Supplementary file 4.** Characteristics of the rehabilitation programmes.

| **Author (year)** | **Early Rehabilitation** | **Delayed Rehabilitation** |
| --- | --- | --- |
| Duzgun, Baltaci (4) | **IP:** NA  **Weeks 0-1:** Cold pack every 2 hours for 20 min  **Weeks 1-2:** Cold pack **+** deltoid and biceps soft-tissue mobilisation **+** shoulder PROM exercises for flexion and abduction **+** elbow and neck AROM **+** hand strengthening  **Weeks 2-3:** Cold pack **+** shoulder PROM exercises for flexion **+** elbow and neck AROM exercises **+** glenohumeral mobilization  **Weeks 3-4:** Cold pack **+** scapular mobilization **+** Shoulder AROM exercises for flexion, internal rotation, abduction **+** strengthening for biceps, triceps and serratus anterior using rubber bands  **Weeks 4-5:** Cold pack **+** shoulder AROM exercises for flexion **+** strengthening of shoulder abduction, internal rotation, external rotation with rubber bands  **Weeks 5-6:** Cold pack **+** progression of shoulder strengthening exercises with more resistant rubber bands **+** posterior capsule stretching  **Weeks 6:** Week 5-6 **+** Resistive PNF exercises  **Weeks 7:** Wall shoulder push-up **+** On-the-table press-up **+** on-the-table push-up | **IP:** NA  **Weeks 0-4:** Early rehabilitation weeks 0-1  **Weeks 4-6:** Early rehabilitation weeks 2-3  **Weeks 6-8:** Early rehabilitation weeks 3-4  **Weeks 8-10:** Early rehabilitation weeks 4-5  **Weeks 10-14:** Early rehabilitation weeks 5-6  **Weeks 14-18:** Early rehabilitation weeks 6  **Weeks 18-22:** Early rehabilitation weeks 7 |
| ***Continue*** |  |  |

**Supplementary file 4.** Characteristics of the rehabilitation programmes.

| **Author (year)** | **Early Rehabilitation** | **Delayed Rehabilitation** |
| --- | --- | --- |
| Duzgun, Baltaci (5) | **IP:** 2 weeks  **Weeks 2-7:** Soft tissue mobilization for the scapulothoracic and glenohumeral joints along with mobilisation exercises (3x week during all weeks).  **Weeks 3:** Shoulder AROM exercises for scaption, flexion and abduction  **Weeks 4:** Light resistive exercises with rubber bands. | **IP:** 4 weeks  **Weeks 4-17:** Soft tissue mobilization for the scapulothoracic and glenohumeral joints along with mobilisation exercises (3x week during all weeks).  **Weeks 6:** Shoulder AROM exercises for scaption, flexion and abduction.  **Weeks 8:** Light resistive exercises with rubber bands. |
| Fawzy, Rizk Mohamed (6) | **IP:** Sling for 6 weeks  **Day 2:** Shoulder PROM exercises for flexion and external rotation + pendulum exercise + elbow, wrist and hand AROM exercises  **Week 6:** Shoulder AROM exercises  **Weeks 12:** Shoulder AROM exercises and strengthening exercises  **From 6 months:** Sports activities  Face-to-face sessions 3x week | **IP:** Sling for 6 weeks  **Week 6:** Shoulder PROM exercises for flexion and external rotation + pendulum + elbow, wrist and hand AROM exercises  **Week 9:** Shoulder AROM exercises  **Weeks 12:** Shoulder AROM exercises + shoulder strengthening exercises  **From 6 months:** Sports activities  Face-to-face sessions 3x week |
| ***Continue*** |  |  |

**Supplementary file 4.** Characteristics of the rehabilitation programmes.

| **Author (year)** | **Early Rehabilitation** | **Delayed Rehabilitation** |
| --- | --- | --- |
| Jenssen, Lundgreen (7) | **IP:** Simple sling for 3 weeks  **Day 1:** Elbow and hand AROM exercises + shoulder PROM exercises + pendulum exercises  **Week 3:** Shoulder AROM exercises (lifting anything greater than the weight of the arm for the first 3 months).  **From 6 months:** Heavy lifting and weight training  Face-to-face sessions 2-3x a week | **IP:** Brace with a small abduction pillow for 6 weeks  **Day 1:** Elbow and hand AROM exercises + shoulder PROM exercises + pendulum exercises  **Week 6:** Shoulder AROM exercises (lifting anything greater than the weight of the arm for the first 3 months).  **From 6 months:** Heavy lifting and weight training  Face-to-face sessions 2-3x a week |
| Keener, Galatz (8) | **IP:** Sling for 6 weeks  **Immediate postoperative:** Pendulum exercise **+** elbow, wrist and hand AROM exercises  **Weeks 1-6:** Shoulder PROM performed by therapist  **Weeks 6-12:** Shoulder AAROM and AROM exercises  **3-4 Months:** Deltoid and scapular stabilizer strengthening  **From 4 months:** Full activities based on patient's progress | **IP:** Sling for 6 weeks  **Immediate postoperative – week 6:** Elbow, wrist and hand AROM exercises; no shoulder mobilization  **Week 6-12:** Shoulder PROM performed by therapist  **3-4 Months:** Shoulder AAROM and AROM exercises  **From 4 months:** Deltoid and scapular stabilizer strengthening; full activities between 5 and 6 months based on patients progress |
| ***Continue*** |  |  |

**Supplementary file 4.** Characteristics of the rehabilitation programmes.

| **Author (year)** | **Early Rehabilitation** | **Delayed Rehabilitation** |
| --- | --- | --- |
| Kim, Chung (9) | **IP:** Brace with abduction pillow (30°) during 4 or 5 weeks  **First day postoperative- week 4/5:** Shoulder PROM exercises for flexion, abduction and external rotation **+** elbow, wrist and hand AROM exercises **+** shrugging of shoulders  **Week 4/5:** Shoulder AAROM exercises  **Week 9/12:** Muscle strengthening  **6 Months:** Return of activities | **IP:** Brace with abduction pillow (30°) during 4 or 5 weeks  **First day postoperative- week 4/5:** Elbow, wrist and hand AROM exercises **+** shrugging of shoulders  **Week 4/5:** Shoulder AROM exercises  **Week 9/12:** Muscle strengthening  **6 Months:** Return of activities |
| Kjær (10) | **IP:** 2 weeks in fixed sling followed by 3 weeks in standard sling  **Weeks 2-5:** Physiotherapist guided PROM exercises + close-chain AAROM and AROM exercises (week 2 only) - AAROM and AROM Restrictions: ABD + FLEX < 90 degrees; IR < 90 degrees in neutral; ER = 0 degrees in neutral  **Weeks 3-5:** Close-chain AAROM and AROM exercises - AAROM and AROM Restrictions: ABD + FLEX < 90 degrees; IR < 90 degrees in neutral; ER < 45 degrees in neutral  **Weeks 6-12:** Therapist-supervised AROM (FLEX, ABD, EXT, ER and IR) with gradually (individually) increased loading and progression from close-chain to open-chain exercises  **Weeks 12-20:** Continuation of rehabilitation in the community  Face-to-face sessions 3x a week | **IP:** 2 weeks in fixed sling followed by 3 weeks in standard sling  **Weeks 2-5:** Physiotherapist guided PROM exercises PROM Restrictions: ABD + FLEX: None IR < 90 degrees in neutral ER < 45 degrees in neutral  **Weeks 6-12:** Therapist-supervised AROM (FLEX, ABD, EXT, ER and IR) with gradually (individually) increased loading and progression from close-chain to open-chain exercises  **Weeks 12-20:** Continuation of rehabilitation in the community  Face-to-face sessions 1x a week |

***Continue***

**Supplementary file 4.** Characteristics of the rehabilitation programmes.

| **Author (year)** | **Early Rehabilitation** | **Delayed Rehabilitation** |
| --- | --- | --- |
| Klintberg, Gunnarsson (11) | **IP:** Sling for 4 weeks  **First day to 4 weeks:** Activation of the rotator cuff + shoulder PROM exercises  **Weeks 4-6 weeks:** Increased loading of the rotator cuff + shoulder AAROM exercises (including in the pool)  **Weeks 6-8:** Shoulder AROM exercises + PROM exercise for shoulder internal rotation  **Weeks 8-10:** Resisted exercises for the rotator cuff + dynamic strengthening exercises for the rotator cuff + scapular muscles through range  **Weeks 10-12:** Water-resisted exercises; eccentric loading of the rotator cuff  **Weeks 12-16:** Eccentric load on the rotator cuff during supervised physiotherapy  **Weeks 16-24:** Dynamic strengthening for rotator cuff and scapular muscles throughout full range + active, water-resisted exercises performed throughout full ROM  **From 24 weeks:** Eccentric load on the rotator cuff during supervised physiotherapy | **IP:** Sling for 6 weeks  **First day to 4 weeks:** Shoulder PROM exercises  **Weeks 6-10:** Shoulder AAROM exercises (at six weeks activation of the rotator cuff)  **Weeks 10-16:** Aquatic AAROM shoulder exercises  **Weeks 16-24:** Strengthening water-resisted exercises + eccentric loading of the rotator cuff  **From 24 weeks:** Eccentric load on the rotator cuff during supervised physiotherapy |
| ***Continue*** |  |  |

**Supplementary file 4.** Characteristics of the rehabilitation programmes.

| **Author (year)** | **Early Rehabilitation** | **Delayed Rehabilitation** |
| --- | --- | --- |
| Koh, Lim (12) | **IP:** Sling with an abduction pillow (20°) during 4 weeks  **Week 5-10:** Shoulder PROM exercises with rope, pulley and cane **+** home-based exercise.  Shoulder AAROM and AROM were allowed as patients obtained nearly full PROM  **Week 11- 6 Months:** Shoulder strengthening using elastic bands  **6 Months:** Return to normal activities, including sports | **IP:** Sling with an abduction pillow (20°) during 8 weeks  **Week 9-14:** Shoulder PROM exercises, pulley and cane **+** home-based exercise. Shoulder AAROM and AROM were allowed as patients obtained nearly full PROM  **Week 15 – 6 Months:** Shoulder strengthening using elastic bands  **6 Months:** Return to normal activities, including sports |
| Lee, Cho (13) | **IP:** Sling with an abduction pillow (30°) during 6 weeks  **First day postoperative – week 6:** Shoulder PROM exercises for flexion and external rotation (2x pd) **+** pendulum exercises **+** shoulder PROM exercises (3x pd) **+** home-based exercises  **Week 6-on:** Shoulder AROM **+** PROM exercises for all directions | **IP:** Sling with an abduction pillow (30°) during 6 weeks  **First day postoperative – week 3:** Self-PROM shoulder flexion **+** CPM (2x pd)  **Week 3-6:** Shoulder PROM exercises (2x pd)  **Week 6-on:** Shoulder strengthening using elastic bands |
| ***Continue*** |  |  |

**Supplementary file 4.** Characteristics of the rehabilitation programmes.

| **Author (year)** | **Early Rehabilitation** | **Delayed Rehabilitation** |
| --- | --- | --- |
| Littlewood, Bateman (14) | **IP:** Advice to remove the sling as soon as possible  **From week 0:** Advice to patients gradually begin to active use of their arms as soon as able within acceptable limits of pain.  One face-to-face session with physiotherapist before hospital discharge and at 2 weeks postoperative.  **After 4 weeks:**  Individualised progression between phases  Further 4 face-to-face sessions  **Phase 1:** Progress to assisted movement then full active movement within pain limits  **Phase 2:** Isometric exercises for all shoulder muscle groups  **Phase 3:** Resisted exercises through range, within limits of pain  **Phase 4:** Functional restoration | **IP:** Sling for 4 weeks  **From week 0:** table slides + elbow, wrist and hand exercises AROM exercises + shoulder PROM exercises for abduction, flexion and lateral rotation movements, within pain limits  One face-to-face session with physiotherapist before hospital discharge and at 2 weeks postoperative.  **After 4 weeks:**  Individualised progression between phases  Further 4 face-to-face sessions  **Phase 1:** Progress to assisted movement then full active movement within pain limits  **Phase 2:** Isometric exercises for all shoulder muscle groups  **Phase 3:** Resisted exercises through range, within limits of pain  **Phase 4:** Functional restoration |
| ***Continue*** |  |  |

**Supplementary file 4.** Characteristics of the rehabilitation programmes.

| **Author (year)** | **Early Rehabilitation** | **Delayed Rehabilitation** |
| --- | --- | --- |
| Mazzocca, Arciero (15) | **IP:** Ultrasling with abduction pillow for 6 weeks  **Day 2 or 3 to week 6:** Shoulder AAROM exercises (external rotation & flexion)  **Week 7:** Shoulder AAROM exercises + stretching  **Week 8-12:** Shoulder AROM exercise + stretching  **Week 13:** Shoulder isometric strengthening  **Week 14:** Shoulder strengthening using theraband for external and internal rotation  **Week 15:** Strengthening using theraband for external and internal rotation, and abduction  **From week 16:** Strengthening using theraband for external and internal rotation, and abduction + PNF  Face-to-face session 2x week | **IP:** Ultrasling with abduction pillow for 6 weeks  **Week 5:** Shoulder AAROM exercises (external rotation & flexion)  **Week 7:** Shoulder AAROM exercises + stretching  **Week 8-12:** Shoulder AROM exercise + stretching  **Week 13:** Isometric strengthening  **Week 14:** Strengthening using theraband for external and internal rotation  **Week 15:** Strengthening using theraband for external and internal rotation, and abduction  **From week 16:** Strengthening using theraband for external and internal rotation, and abduction + PNF  Face-to-face session 2x week |
| Oyarzún, Poblete (16) | **IP:** Sling + abduction component for 1 week  **Weeks 2-4:** Shoulder PROM exercises for flexion & external rotation + shoulder AROM exercise for adduction and trunk rotations + isometric strengthening for shoulder internal rotation and hand gripping + strengthening exercises for elbow flexion  Face-to-face sessions 4x week for 4 weeks | **IP:** Sling + abduction component for 4 weeks  **Weeks 2-4:** Pendulum exercise 2x day for 4x week |
| ***Continue*** |  |  |

**Supplementary file 4.** Characteristics of the rehabilitation programmes.

| **Author (year)** | **Early Rehabilitation** | **Delayed Rehabilitation** |
| --- | --- | --- |
| Raschhofer, Poulios (17) | **IP:** Sling for 6 weeks  **Week 1:** Elbow and shoulder girdle AROM exercises + shoulder PROM exercise elevation (45°) and abduction (45°)  **Weeks 2-6:** Isometric strengthening  **Week 6-onwards:** dynamic activation of the rotator cuff and strengthening of shoulder muscles  20 sessions in 12 weeks | **IP:** Sling for 6 weeks  **Week 1:** Elbow and shoulder girdle AROM exercises + shoulder PROM exercise elevation (45°) and abduction (45°)  **Weeks 2-6:** no information  **Week 6-onwards:** dynamic activation of the rotator cuff and strengthening of shoulder muscles  20 sessions in 12 weeks |
| Sheps, Bouliane (18) | **IP:** Sling as needed  Shoulder PROM, AAROM and AROM exercises for ADLs as early as pain allowed.  After 6 weeks the rehabilitation programme was identical between groups. No further information was available for the rehabilitation programme from 6 weeks postoperative | **IP:** Sling for 6 weeks Shoulder PROM and AAROM were allowed No active movements of the shoulder. After 6 weeks the rehabilitation programme was identical between groups. No further information was available for the rehabilitation programme from 6 weeks postoperative |
| ***Continue*** |  |  |

**Supplementary file 4 (continue).** Characteristics of the rehabilitation programmes.

| **Author (year)** | **Early Rehabilitation** | **Delayed Rehabilitation** |
| --- | --- | --- |
| Sheps, Silveira (19) | **IP:** Sling only for comfort  **Weeks 0-6:** Pendulum exercises + shoulder AAROM as pain allows + elbow and hand AROM exercises + pain-free AROM  To begin general conditioning program of choice  **Weeks 6-10:** Shoulder AROM exercises as pain allows – all planes + gentle stretching into terminal ROM + initiate closed chain exercises.  **Weeks 10-26:** Isometric strengthening; progress to isotonic strengthening within pain-free ROM  Closed chain strengthening  Overhead strengthening once full ROM achieved and pain well controlled.  Should not lift >15 lb. unless specified by physician  Continue with stretching (therapist may now assist)  Joint mobilization permitted | **IP:** Sling for 6 weeks **Weeks 0-6:** Pendulum exercises + shoulder AAROM were allowed + elbow and hand AROM exercises No active movements of the shoulder  To begin general conditioning program of choice  **Weeks 6 -10:** Shoulder AROM exercises as pain allows – all planes + gentle stretching into terminal ROM + initiate closed chain exercises.  **Weeks 10-26:** Isometric strengthening; progress to isotonic strengthening within pain-free ROM  Closed chain strengthening  Overhead strengthening once full ROM achieved and pain well controlled.  Should not lift >15 lb. unless specified by physician  Continue with stretching (therapist may now assist)  Joint mobilization permitted |
| ***Continue*** |  |  |

**Supplementary file 4 (continue).** Characteristics of the rehabilitation programmes.

| **Author (year)** | **Early Rehabilitation** | **Delayed Rehabilitation** |
| --- | --- | --- |
| Tirefort, Schwitzguebel (20) | **IP:** No sling  **Week 0-4:** Shoulder PROM and AAROM exercises  **From 4 weeks:** Progressive shoulder AROM exercises with elbow at the side. No exercises involving lifting of the elbow in any direction, unless assisted, were permitted  **From 8 weeks:** Demanding activities and light sports  **From 12 weeks:** Shoulder strengthening | **IP:** Sling for 4 weeks  **Week 0-4:** Shoulder PROM and AAROM exercises  **From 4 weeks:** Progressive shoulder AROM exercises with elbow at the side. No exercises involving lifting of the elbow in any direction, unless assisted, were permitted  **From 8 weeks:** Demanding activities and light sports  **From 12 weeks:** Shoulder strengthening |

AAROM: Active Assisted Range of movement, ABD: Abduction, ADLs: Activities of Daily Living, CPM: Continuous Passive Motion, ER: External Rotation, FLEX: Flexion, GH: glenohumeral, IP: Immobilisation Period, IR: Internal Rotation, NA: Not Available, PNF: Proprioceptive neuromuscular facilitation, PROM: Passive Range of Movement, ROM: Range Of Movement.

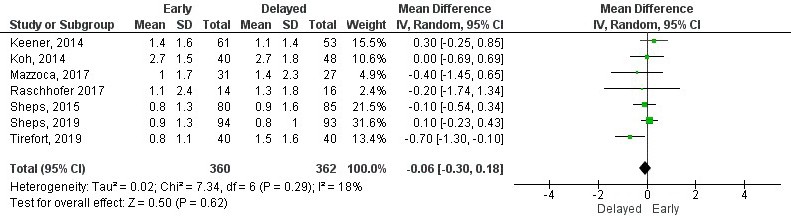
**Supplementary file 5.** Forest plots.

**Supplementary file 5.1**. Forest plot of pain intensity at six months by visual analogue scale.

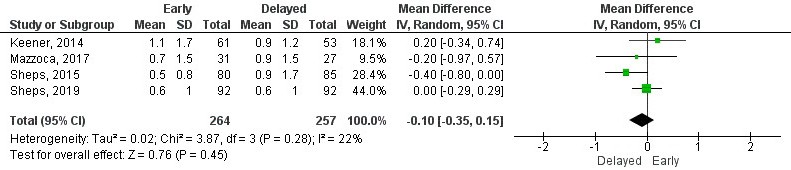

**Supplementary file 5.2**. Forest plot of pain intensity at one year by visual analogue scale.

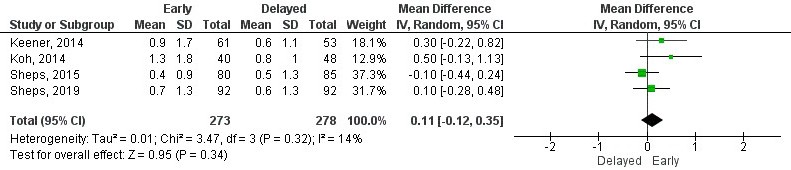
**Supplementary file 5.3**. Forest plot of pain intensity at two years by visual analogue scale.

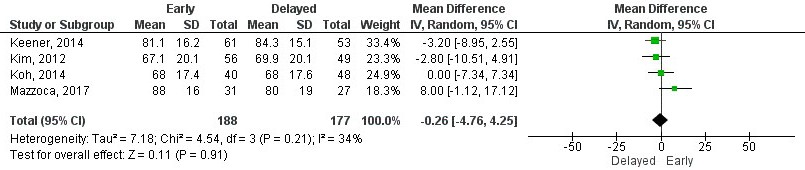

**Supplementary file 5.4**. Forest plot of function at six months by American Shoulder and Elbow Surgery score.

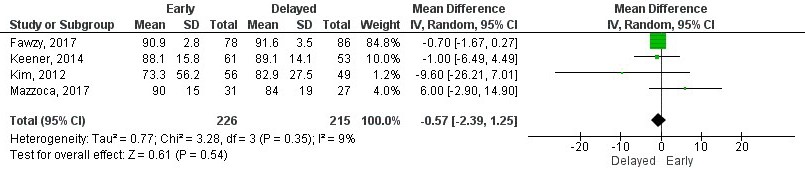
**Supplementary file 5.5**. Forest plot of function at one year by American Shoulder and Elbow Surgery score.

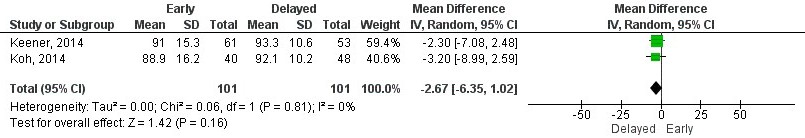

**Supplementary file 5.6**. Forest plot of function at two years by American Shoulder and Elbow Surgery score.

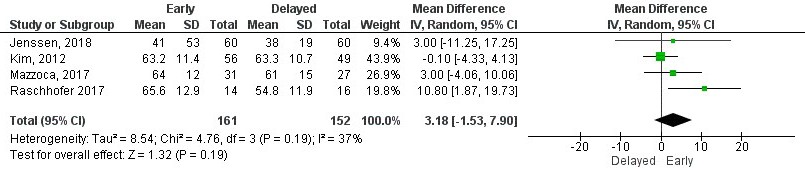
**Supplementary file 5.7**. Forest plot of function at three months years by Constant-Murley score.

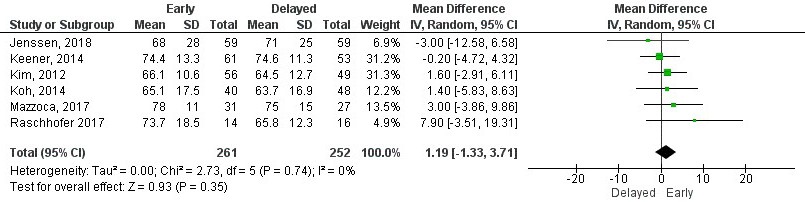

**Supplementary file 5.8**. Forest plot of function at six months by Constant-Murley score.

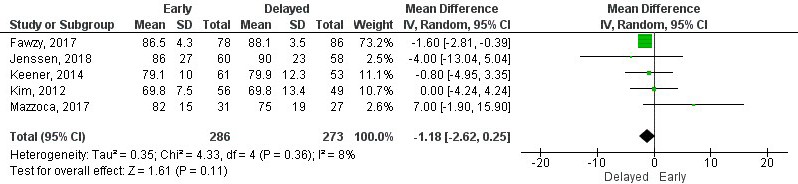
**Supplementary file 5.9**. Forest plot of function at one year by Constant-Murley score.

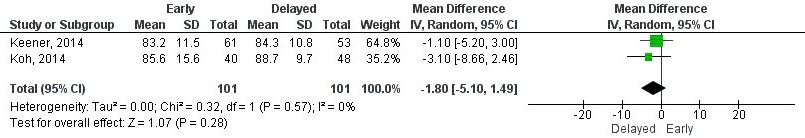

**Supplementary file 5.10**. Forest plot of function at two years by Constant-Murley score.

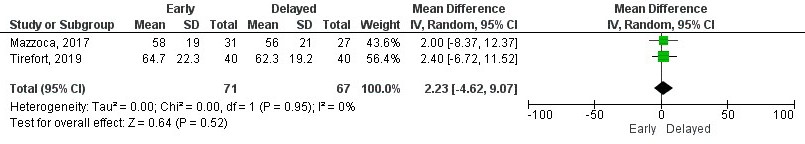
**Supplementary file 5.11**. Forest plot of function at three months by Single Assessment Numeric Evaluation.

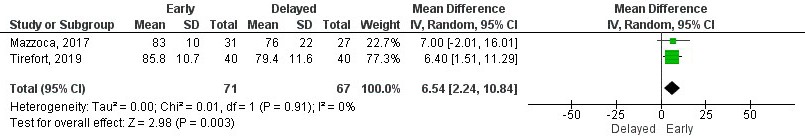

**Supplementary file 5.12**. Forest plot of function at six months by Single Assessment Numeric Evaluation.

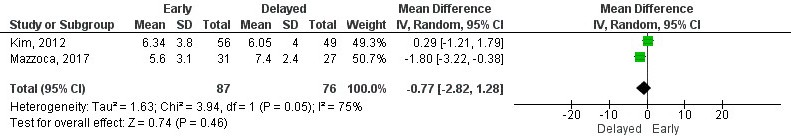
**Supplementary file 5.13**. Forest plot of function at three months by Simple Shoulder Test.

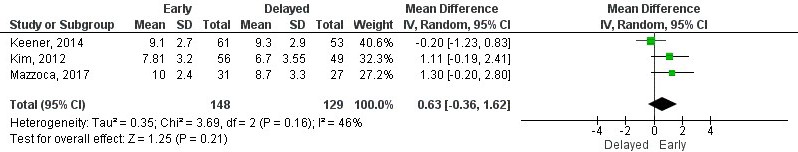

**Supplementary file 5.14**. Forest plot of function at six months by Simple Shoulder Test.

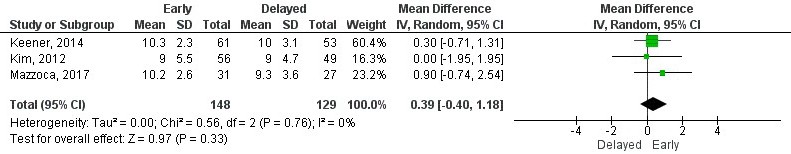
**Supplementary file 5.15**. Forest plot of function at one year by Simple Shoulder Test.

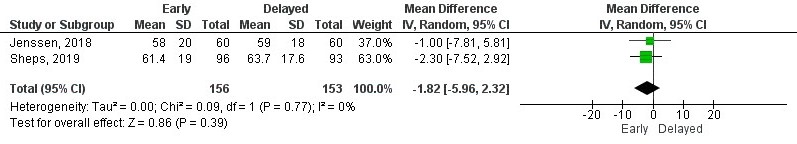

**Supplementary file 5.16.** Forest plot of function at three months by Western Ontario Rotator Cuff Index.

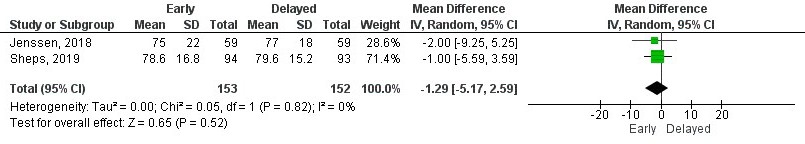
**Supplementary file 5.17.** Forest plot of function at six months by Western Ontario Rotator Cuff Index.

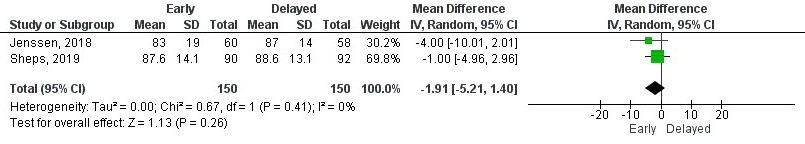

**Supplementary file 5.18.** Forest plot of function at one year by Western Ontario Rotator Cuff Index.

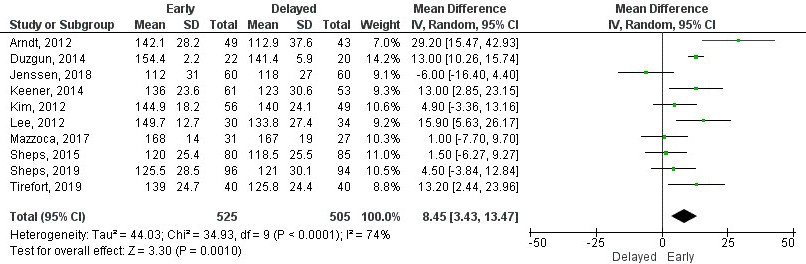

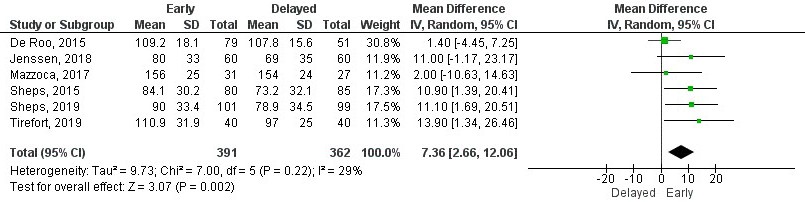
**Supplementary file 5.19.** Forest plot of range of movement for shoulder flexion at six weeks.

**Supplementary file 5.20.** Forest plot of range of movement for shoulder flexion at three months.

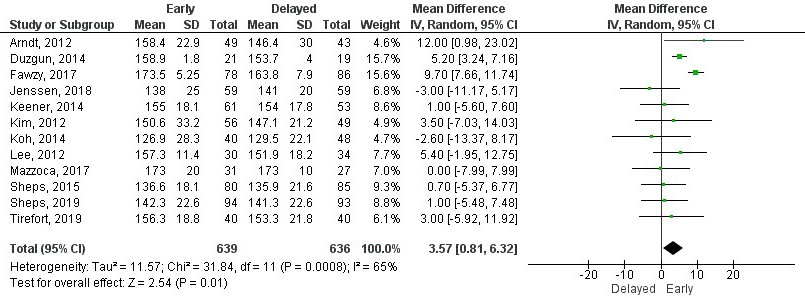
**Supplementary file 5.21.** Forest plot of range of movement for shoulder flexion at six months.

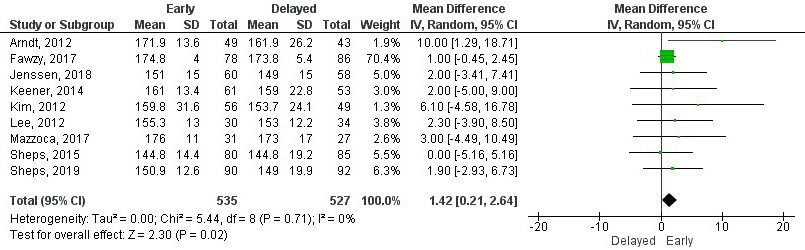

**Supplementary file 5.22.** Forest plot of range of movement for shoulder flexion at one year.

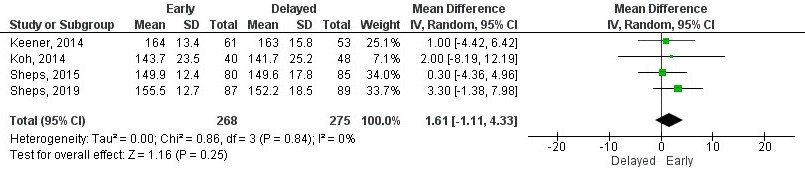
**Supplementary file 5.23.** Forest plot of range of movement for shoulder flexion at two years.

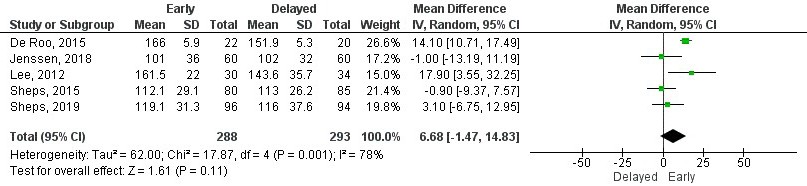

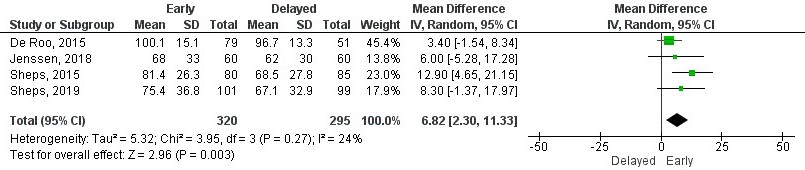
**Supplementary file 5.24.** Forest plot of range of movement for shoulder abduction at six weeks.

**Supplementary file 5.25.** Forest plot of range of movement for shoulder abduction at three months.

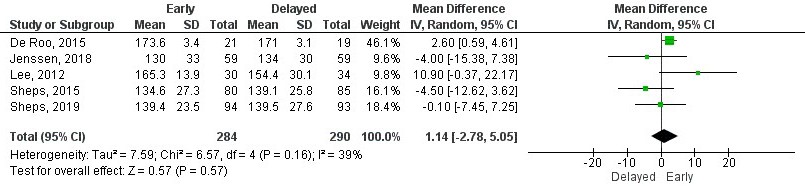
**Supplementary file 5.26.** Forest plot of range of movement for shoulder abduction at six months.

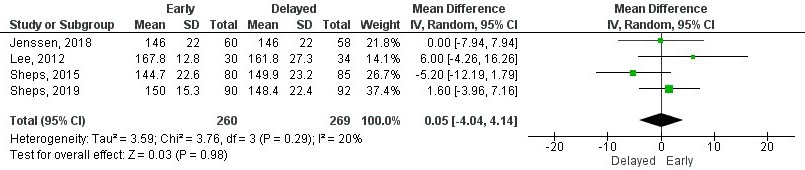

**Supplementary file 5.27.** Forest plot of range of movement for shoulder abduction at one year.

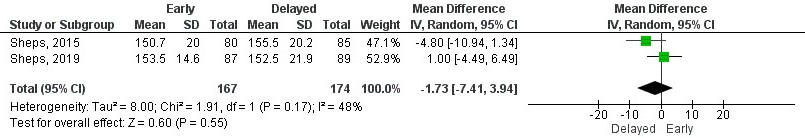

**Supplementary file 5.28.** Forest plot of range of movement for shoulder abduction at two years.

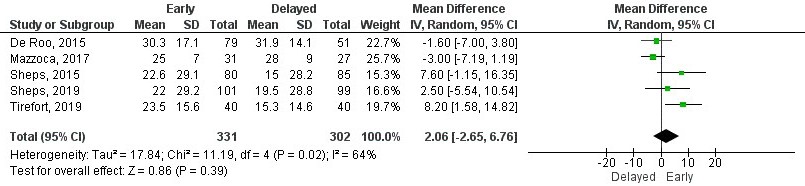

**Supplementary file 5.29.** Forest plot of range of movement for external rotation at six weeks.

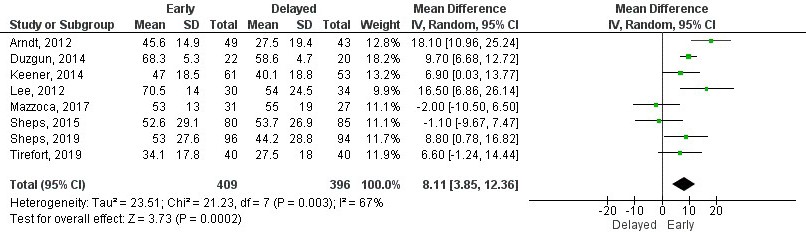
**Supplementary file 5.30.** Forest plot of range of movement for external rotation at three months.

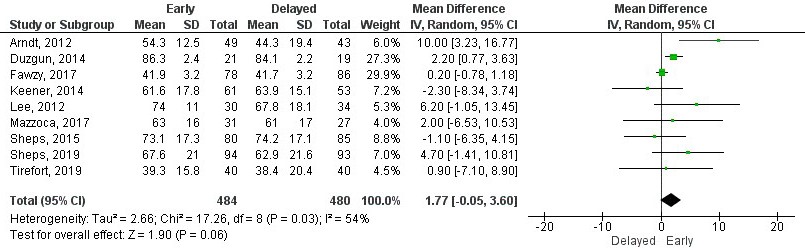

**Supplementary file 5.31.** Forest plot of range of movement for external rotation at six months.

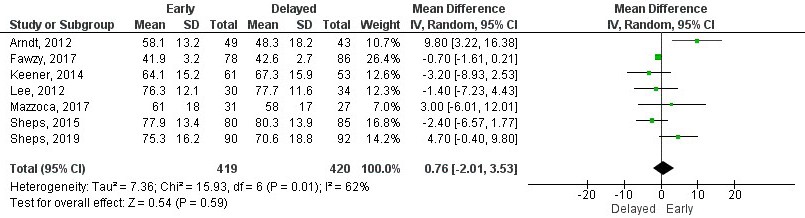
**Supplementary file 5.32.** Forest plot of range of movement for external rotation at one year.

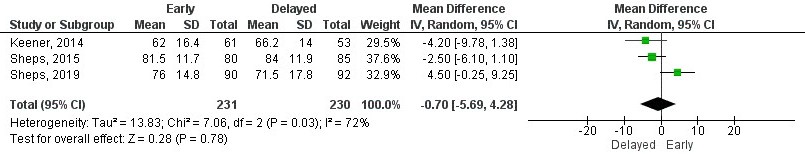

**Supplementary file 5.33.** Forest plot of range of movement for external rotation at two years.

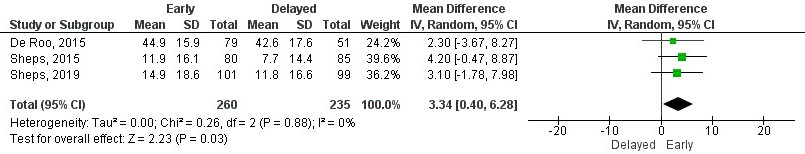
**Supplementary file 5.34.** Forest plot of range of movement for internal rotation at six weeks.

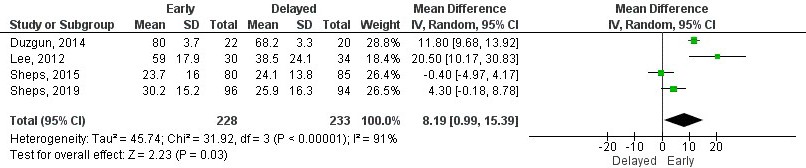

**Supplementary file 5.35.** Forest plot of range of movement for internal rotation at three months.

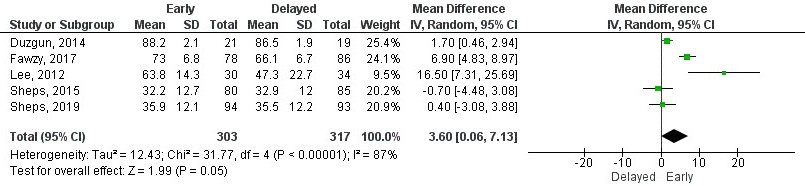

**Supplementary file 5.36.** Forest plot of range of movement for internal rotation at six months.

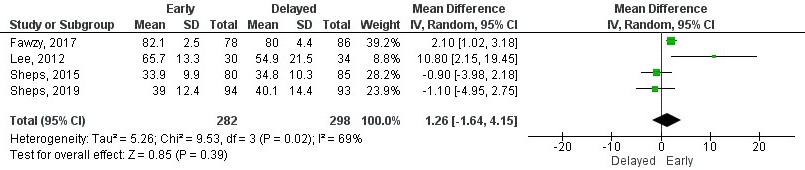

**Supplementary file 5.37.** Forest plot of range of movement for internal rotation at one year.

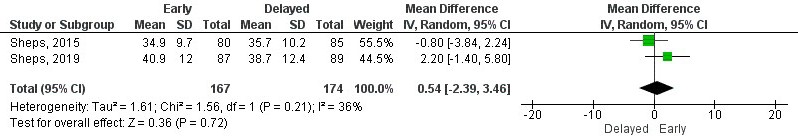

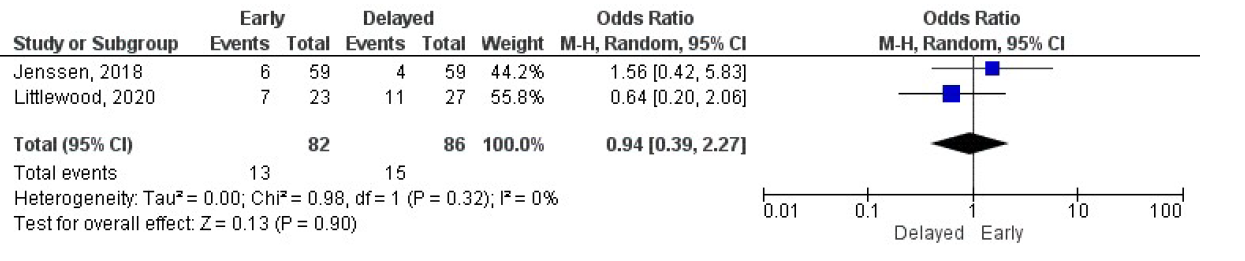
**Supplementary file 5.38.** Forest plot of range of movement for internal rotation at two years.

**Supplementary file 5.39.** Forest plot of odds ratio for repair integrity at three months.

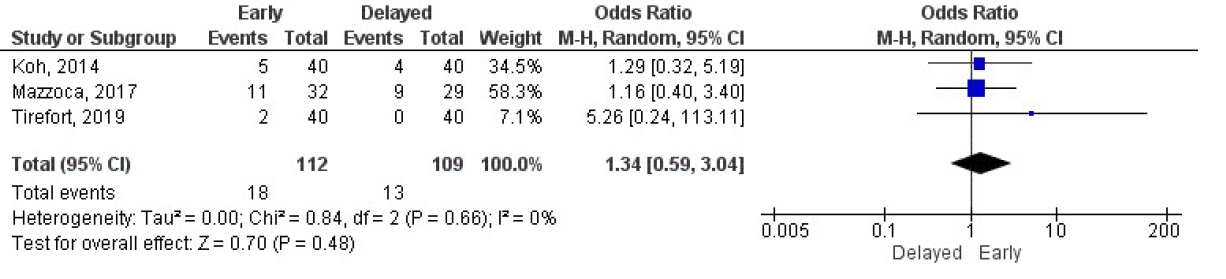

**Supplementary file 5.40.** Forest plot of odds ratio for repair integrity at six months.

**Supplementary file 6.** GRADE summary of findings.

| **Outcomes** | **№ of participants  (studies) Follow up** | **Certainty of the evidence (GRADE)** | **Relative effect (95% CI)** |
| --- | --- | --- | --- |
| Pain - Visual Analogue Scale at 6 weeks | 707 (6 RCTs) | ⨁⨁◯◯ LOW ^a,b^ | - |
| Pain - Visual Analogue Scale at 3 months | 692 (6 RCTs) | ⨁⨁⨁⨁ HIGH | - |
| Pain - Visual Analogue Scale at 6 months | 722 (7 RCTs) | ⨁⨁⨁⨁ HIGH | - |
| Pain - Visual Analogue Scale at 1 year | 521 (4 RCTs) | ⨁⨁⨁⨁ HIGH | - |
| Pain - Visual Analogue Scale at 2 years | 551 (4 RCTs) | ⨁⨁⨁⨁ HIGH | - |
| Function - American Shoulder and Elbow Score at 3 months | 243 (3 RCTs) | ⨁⨁⨁◯ MODERATE ^c^ | - |
| Function - American Shoulder and Elbow Score at 6 months | 365 (4 RCTs) | ⨁⨁⨁◯ MODERATE ^c^ | - |
| Function - American Shoulder and Elbow Score at 12 months | 441 (4 RCTs) | ⨁⨁◯◯ LOW ^d,e^ | - |
| Function - American Shoulder and Elbow Score at 24 months | 202 (2 RCTs) | ⨁⨁⨁◯ MODERATE ^c^ | - |
| Function - Constant-Murley score at 3 months | 313 (4 RCTs) | ⨁⨁◯◯ LOW ^c,e^ | - |
| Function - Constant-Murley score at 6 months | 513 (6 RCTs) | ⨁⨁⨁⨁ HIGH | - |
| Function Constant-Murley score at 12 months | 559 (5 RCTs) | ⨁⨁◯◯ LOW ^d,e^ | - |
| Function - Constant-Murley score at 24 months | 202 (2 RCTs) | ⨁⨁⨁◯ MODERATE ^c^ | - |
| Function - Single Assessment Numeric Evaluation at 3 months | 138 (2 RCTs) | ⨁⨁⨁◯ MODERATE ^c^ | - |
| Function - Single Assessment Numeric Evaluation at 6 months | 138 (2 RCTs) | ⨁⨁⨁◯ MODERATE ^c^ | - |
| Function - Simpe Shoulder Test at 3 months | 163 (2 RCTs) | ⨁◯◯◯ VERY LOW ^a,b,c,d^ | - |
| Function - Simple Shoulder Test at 6 months | 277 (3 RCTs) | ⨁⨁⨁◯ MODERATE ^c^ | - |
| Function - Simple Shoulder Test at 12 months | 277 (3 RCTs) | ⨁⨁⨁◯ MODERATE ^c^ | - |
| Function - Western Ontario Rotator Cuff Index at 3 months | 309 (2 RCTs) | ⨁⨁⨁◯ MODERATE ^c^ | - |
| Function - Western Ontario Rotator Cuff Index at 6 months | 363 (2 RCTs) | ⨁⨁⨁◯ MODERATE ^c^ | - |
| Function - Western Ontario Rotator Cuff Index at 12 months | 300 (2 RCTs) | ⨁⨁⨁◯ MODERATE ^c^ | - |
| Range of movement - Flexion at 6 weeks | 753 (6 RCTs) | ⨁⨁⨁⨁ HIGH | - |
| Range of movement - Flexion at 3 months | 1030 (10 RCTs) | ⨁⨁◯◯ LOW ^a,d^ | - |
| Range of movement - Flexion at 6 months | 1275 (12 RCTs) | ⨁⨁◯◯ LOW ^a,d^ | - |
| Range of Movement - Flexion at 12 months | 898 (9 RCTs) | ⨁⨁⨁◯ MODERATE ^d^ | - |
| Range of Movement - Flexion at 24 months | 543 (4 RCTs) | ⨁⨁⨁⨁ HIGH | - |
| Range of Movement - Abduction at 6 weeks | 615 (4 RCTs) | ⨁⨁⨁◯ MODERATE ^f^ | - |
| Range of Movement - Abduction at 3 months | 581 (5 RCTs) | ⨁⨁◯◯ LOW ^a,b,e,f^ | - |
| Range of Movement - Abduction at 6 months | 574 (5 RCTs) | ⨁⨁◯◯ LOW ^e,f^ | - |
| Range of Movement - Abduction at 12 months | 529 (4 RCTs) | ⨁⨁⨁◯ MODERATE ^e^ | - |
| Range of Movement - Abduction at 24 months | 341 (2 RCTs) | ⨁⨁⨁◯ MODERATE ^c^ | - |
| Range of Movement - External Rotation at 6 weeks | 633 (5 RCTs) | ⨁⨁⨁◯ MODERATE ^a^ | - |
| Range of Movement - External Rotation at 3 months | 805 (8 RCTs) | ⨁⨁⨁◯ MODERATE ^a^ | - |
| Range of Movement - External Rotation at 6 months | 964 (9 RCTs) | ⨁⨁◯◯ LOW ^a,d^ | - |
| Range of Movement - External Rotation at 12 months | 839 (7 RCTs) | ⨁⨁◯◯ LOW ^a,d^ | - |
| Range of Movement - External Rotation at 24 months | 461 (3 RCTs) | ⨁⨁⨁◯ MODERATE ^a^ | - |
| Range of Movement - Internal Rotation at 6 weeks | 495 (3 RCTs) | ⨁⨁⨁⨁ HIGH | - |
| Range of Movement - Internal Rotation at 3 months | 461 (4 RCTs) | ⨁◯◯◯ VERY LOW ^a,b,d,e^ | - |
| Range of Movement - Internal Rotation at 6 months | 620 (5 RCTs) | ⨁⨁◯◯ LOW ^a,d,e^ | - |
| Range of Movement - Internal Rotation at 12 months | 580 (4 RCTs) | ⨁⨁◯◯ LOW ^a,d,e^ | - |
| Range of Movement - Internal Rotation at 24 months | 341 (2 RCTs) | ⨁⨁⨁◯ MODERATE ^c^ | - |
| Repair integrity at 3 months | 168 (2 RCTs) | ⨁⨁⨁◯ MODERATE ^g^ | **OR 0.94** (0.39 to 2.27) |
| Repair integrity at 6 months | 221 (3 RCTs) | ⨁⨁⨁◯ MODERATE ^h^ | **OR 1.34** (0.59 to 3.04) |
| Repair integrity at 12 months | 960 (8 RCTs) | ⨁⨁◯◯ LOW ^d,i^ | **OR 1.26** (0.82 to 1.93) |

***The risk in the intervention group** (and its 95% confidence interval) is based on the assumed risk in the comparison group and the **relative effect** of the intervention (and its 95% CI).
**CI:** Confidence interval; **MD:** Mean difference; **OR:** Odds ratio

a. High heterogeneity

b. Large confidence interval. Confidence interval included both the point of no effect (0) and the minimal clinically important difference (MCID) for each outcome

MCID values:

- Pain: 1.4

- American Shoulder and Elbow Surgery: 11.1

- Constant Murley: 10.4

- Single Assessment Numeric Evaluation: 16.9

- Simple Shoulder Test: 2.3

- Western Ontario Rotator Cuff: 245

- Flexion range of movement: 14

- Abduction range of movement: 7

- Internal and external rotation range of momvent:15

c. Sample size <400 (200 per group)

d. A substantial number of trials were rated as of unclear and/or high risk of bias

e. Wide variance of point estimates across studies

f. Trial with a high risk of bias receiving most of the weight in the analysis

g. Small sample size (optimal information size: 8614 per group)

h. Small sample size (optimal information size: 1096 per group)

i. Small sample size (optimal information size: 3198 per group)
